# Guiding the development of climate counterfactuals for health impact attribution studies

**DOI:** 10.64898/2026.06.16.26355779

**Authors:** Gina E C Charnley, Maximilian Kotz, Ania Kawiecki, Katherine M Grayson

## Abstract

Climate change detection and attribution (D&A) methods have become vital for quantifying the influence of anthropogenic forcing on the Earth’s systems, including human health. Health impact attribution (HIA) studies seek to disentangle climate-driven health effects from natural variability yet are often constrained by the availability of accessible counterfactual climate scenarios. This tutorial paper presents a flexible, reproducible framework for developing counterfactual climates without reliance on computationally intensive global circulation models. We provide practical, R-based methodologies for constructing both trend-based (temperature and non-temperature) and event-based counterfactual, using a variety of techniques including model residual detrending, data-driven decomposition (e.g., Singular Spectrum Analysis and Empirical Mode Decomposition) and stochastic weather generators. The tutorial also explores the incorporation of greenhouse gas concentrations as forcing variables, rather than global mean temperature anomalies. By operationalising these methods through worked examples and an open code repository, this paper aims to build capacity within the HIA community, enhance methodological transparency, and foster interdisciplinary collaboration between climate and health researchers.

## Introduction

Climate change is defined as a change in the state of the climate that can be identified by changes in the mean and/or variability of its properties, that persist over decades or longer. Across geological and interannual timescales, natural forcings have always driven climate change; however, the dominant cause of climate change observed since the mid-20th century has been anthropogenic rather than natural, with natural processes more often contributing to shorter-term climate variability. Human activity relates to the release of greenhouse gases (GHG) (e.g., CO_2_, CH_4_, N_2_O, O_3_ and H_2_O) into the atmosphere through burning of fossil fuels, land use change, and industrial or commercial processes.^1,2^

Climate-related detection and attribution (D&A) methods are a set of methodological approaches designed to disentangle the role of human forcing in climate dynamics. The field of D&A grapples with detecting changes in the Earth’s climate, distinguishing these changes from natural internal variability, attributing these changes to various forcings and understanding how today’s earth system would be different in the absence of anthropogenic climate change. The goal of D&A methodologies is primarily to advance our understanding of the role of anthropogenic forcing on the climate, by comparing possible alternative worlds without this forcing, known as counterfactual climates.^2,3^ A counterfactual concerns the science of what did not happen and relates to “what-if” reasoning. It has deep roots in mathematics, logic, and physics, long before becoming widely used in causal inference and philosophy.^10^

Climate change poses risks to many sectors, including but not limited to agriculture, urban planning, energy, forestry, fishing and tourism.^4^ Due to the wide-ranging impacts of climate change, several fields of scientific research (including D&A) have emerged in climate-related impact sectors. A rapidly expanding field of study is climate-related health impacts, including communicable and non-communicable diseases, temperature-related mortality and air quality and pollution.^42–45^ Due to the growing evidence that climate and weather and therefore climate change can pose a threat to human health, detecting and attributing the burden of human health caused by anthropogenic climate change, termed health impact attribution (HIA), has become an emerging interdisciplinary field in the last decade. The first HIA study is reported to have been in 2016, and since then there have been several published studies especially on temperature-related mortality.^5,6^

HIA uses a wide range of methodologies, often depending on the health outcome of interest, the climate/weather variables, the availability of a counterfactual climate, the objectives of the study and how the results are to be communicated. These approaches can be broadly categorised within two distinct branches; trend-based and event-based. Trend-based methodologies explore the impact of long-term climate changes on a health outcome whereas event-based approaches investigate a specific or multiple extreme weather event(s) and how this/they impacted a health outcome.^7^ Figure 1 outlines a HIA workflow and how counterfactual climate variables are used in this context. The first step is to outline the attribution question e.g., what would health outcome X have been in the absence of hazard Y/anthropogenic climate change. The second would be to specify and fit an exposure-response model, by harmonising the data and selecting a study design and modelling framework. Once the model is fit and the performance is satisfactory, one can generate counterfactual exposures and use the health model to make factual and counterfactual predictions (for current climatic conditions and a climate without anthropogenic forcings, respectively). Finally, one can compute the attributable impacts, which could be expressed as counts (e.g., number of cases/deaths) or as a fraction.

**Figure 1:**
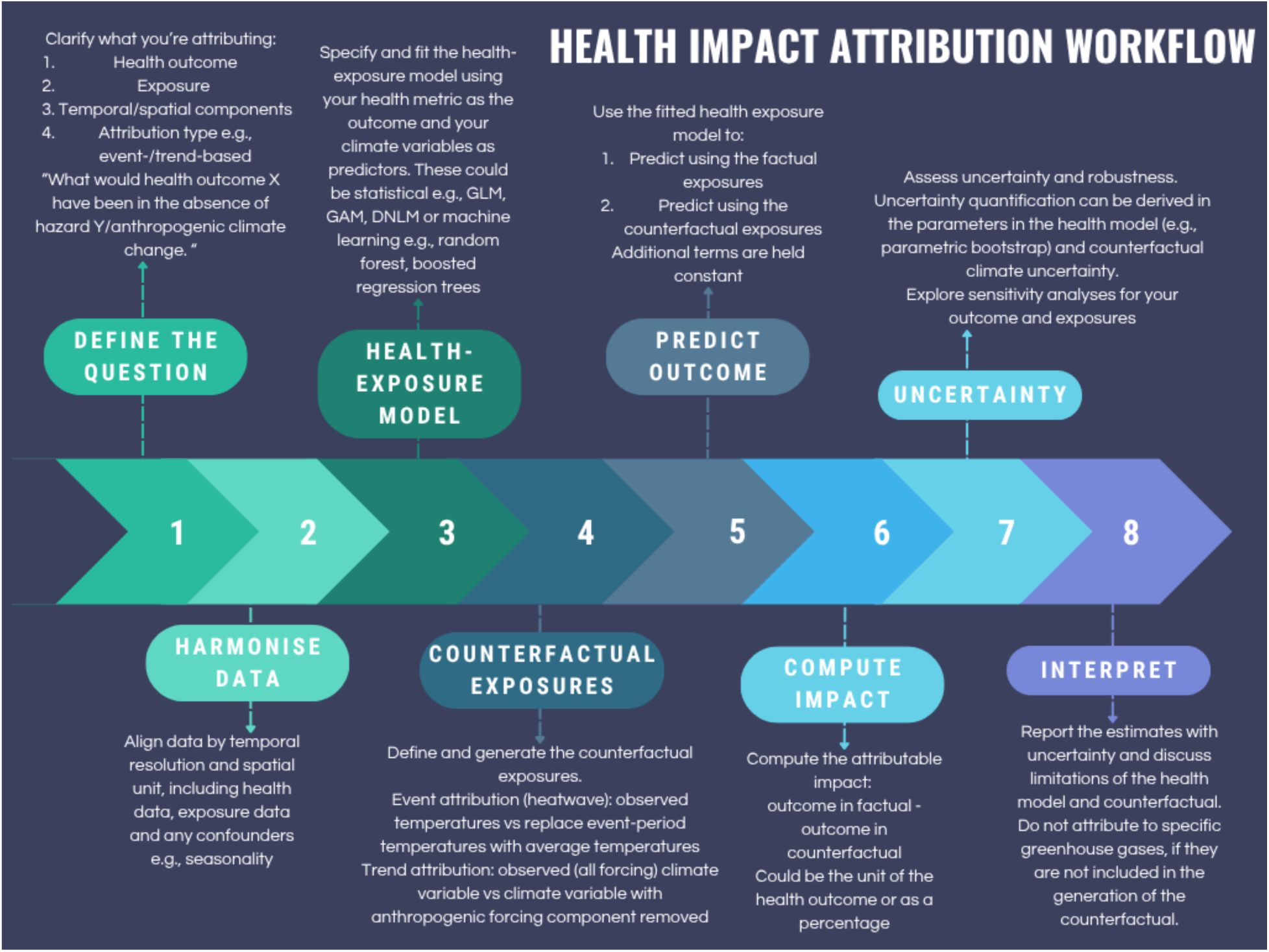
Demonstration of a health impact attribution workflow, covering eight key steps from defining the attribution question to interpretation and reporting. Acronyms include generalised linear model (GLM), generalised additive model (GAM), distributed lag non-linear model (DLNM). *Source*: Author’s own

As HIA is a new and evolving field, methodologies are improving, however, current limitations include an assumption of a static exposure-response relationship and a lack of incorporation of socio-demographic variables which are often more influential for health than climate.^9^ HIA is also constrained by the inherent challenges of generating counterfactual climates. These include difficulties in isolating the forced climate response from a relatively short observational record, high internal variability in certain variables such as precipitation, and limited agreement among General Circulation Models (GCMs) for some climate components.^8^ GCMs at regional or global scale are complex and computationally intensive, making them less interpretable and accessible across disciplines. Furthermore, counterfactual climate variables are ideally needed for all variables included in the health model; while it is feasible to perturb only a subset of variables (e.g., temperature), this restricts attribution to specific climate-mediated pathways rather than capturing the full effect of anthropogenic climate change on health outcomes.^11^ Pre-existing, open-source outputs from GCMs, such as ISIMIP and DAMIP, are readily available online and can be used as counterfactuals. However, these are generated for a specific set of climate variables over defined spatio-temporal resolutions, which may not match what is used in the health-exposure model.^12,13^

Here, we aim to explore the emerging field of data-driven methodologies for climate counterfactuals, a set of computationally inexpensive and flexible approaches which are accessible to a wide range of end users across different fields. We will guide the user through worked examples in R to enhance their knowledge of climate change and the development of counterfactual climates, deepening understanding and hopefully leading to better interpretation of results. The methodologies presented will include a broad range including both trend-based and event-based counterfactuals, detrending methods using temperature and non-temperature variables and the incorporation of GHG concentration data. Although this is not an exhaustive list of methods, we believe that the set of methodologies presented are enough to establish counterfactuals that cover the current scope of HIA research. This tutorial builds on the work of the seminal ATTRICI methodology,^14^ and the publication of its methods and datasets. In combination with the work presented here, a GitHub repository is available to complement this tutorial (see Code and Data Availability).

## Data generation and processing

We generated several synthetic climate datasets representing longer-term monthly temperature, precipitation, and relative humidity (1920–2025), as well as a shorter daily temperature dataset with embedded heatwaves (2010–2019), hereafter referred to as heatwave data. We generated our own data, rather than using real-world climate data to keep the file size small and computationally un-intensive. These data are generated and saved in a csv format in the file “02_synthetic_climate_data_generation” in the GitHub repository.

The long-term monthly values were constructed for a spatial domain covering the Delhi region on a 1° grid (28.5–29.5°N, 76.5–77.5°E). The simulations aimed to capture realistic climatological behaviour including trends, seasonality, spatial variation, and autocorrelated noise. Temperature at each grid cell and month was modelled as the combination of four components (saved as “temperature.csv”):

1. A long-term warming trend – A linear trend was applied corresponding to ∼1.2 °C warming per century, scaled to monthly resolution.
2. A seasonal cycle – Annual temperature seasonality was imposed using a sinusoidal function with a 10 °C amplitude.
3. Autocorrelated stochastic variability – Month-to-month noise followed an AR(1) process (ϕ = 0.6) to incorporate realistic temporal persistence.
4. Spatial temperature structure – A spatially varying baseline temperature field was generated to approximate Delhi’s climatology (20–35 °C range), including weak latitudinal gradients, a modest cool anomaly in the centre of the domain, and small-scale random variation.

Synthetic monthly precipitation and relative humidity were generated over the same domain and time-period using independent random seeds. Precipitation (mm month⁻¹) combined: a baseline mean of 80 mm, a small increasing long-term trend (1% per decade), a monsoon-shaped seasonal cycle represented by a shifted sine wave (peak in July), stochastic variability incorporated via an AR(1) process (ϕ = 0.5, SD = 20 mm). Negative values were truncated at zero. Humidity (%) incorporated: a baseline of 75% (and restricted to the plausible range of 20-100%), a weak long-term declining trend (−0.5% per century), seasonal variability (amplitude 8%, inverted relative to temperature), AR(1) noise (ϕ = 0.4, SD = 2%). Both variables were merged with the spatial grid and exported as “precipitation.csv” and ‘humidity.csv”.

For the heatwaves data, daily baseline temperatures were constructed (not for a specific location) using: a mean temperature of 15 °C, annual and semi-annual seasonal harmonics to reproduce realistic intra-annual variability, a modest warming trend (∼0.4°C over the decade), and gaussian noise (SD = 2 °C). Artificial heatwaves were then embedded to represent extreme hot events, 1–3 synthetic heatwaves per year were inserted during June–August. characterised by a random start date, a duration of 5–14 days, a smooth temperature anomaly peaking at +5 to +12°C. Heatwaves were imposed using a sinusoidal anomaly to avoid unrealistic step changes. Heatwave detection was identified following a standard exceedance-based definition of temperatures exceeding the 90th percentile for ≥3 consecutive days and were flagged accordingly. In order to explore the use of empirical relationships in counterfactual climates, we generated some complementary observational data for other climate variables relative to the temperatures (precipitation, water vapour and convective available potential energy (CAPE)) in the heatwave dataset following known relationships e.g., water vapour increased with temperature, heatwaves suppressed rainfall, extreme precipitation generated as random and rare (2% chance) and independent of normal precipitation and CAPE correlated to temperature and humidity. The resulting daily dataset was saved as “synthetic_heatwaves.csv”.

The raw Berkeley Earth global mean temperature anomalies (GMTA) data were downloaded for use here.^19^ These data are monthly to a 1×1 degree grid cell and compute temperature anomalies using 1850-1900 as the “pre-industrial” reference period. The data have been converted to annual to remove the seasonal component, using a weighted mean to account for different month length and saved in a csv format. The data and processing script are saved in the file “03_gmta_processing”.

Finally, we explore the use of greenhouse gas (GHG) concentration data, as a substitute for GMTA in the detrending process, as these two variables are highly correlated and can allow the user to attribute the health outcome to GHGs, rather than to climate change more generally. Here, we use a combination of the NOAA Mauna Loa record and the Law Dome Ice Cores, which are then joined to provide a long-term dataset of GHG concentrations.^24,25^ The raw, cleaned data in a csv format and processing script are saved in the file “04_ghg_concentrations_processing”.

## Counterfactual climate trends

The aim of creating a trend counterfactual is to remove the anthropogenic forcing component from a long-term time-series of a climate variable, they are computationally unexpensive compared to GCM generated counterfactuals and allow for more flexibility over temporal and spatial scales. Detrending is a highly interpretable method often used to generate trend counterfactuals, it is based on the assumption that the most recent warming is due to anthropogenic climate change and is particularly useful when working with observation datasets, rather than model simulations.^15^ Detrending can be achieved via empirical approaches that regress local climate conditions on GMTA and use the residuals as a counterfactual in the absence of human-forced climate change, as used in the ATTRICI method.^14^ GMTA are generally used to extract the forced component for temperature counterfactuals, however, they can be used in the same way for other climate variable counterfactuals (to represent the forced component).^14^ Another method of detrending includes the use of data-driven decomposition methods (e.g., Singular Spectrum Analysis (SSA), Empirical Mode Decomposition (EMD), wavelet detrending and differencing), which separate long-term trends from variability allowing the construction of a counterfactual by removing the trend component (which we assume is the forced component). These methods can be used on any climate variable; however, they are more computationally intensive than residual-based approaches and require a clear trend to decompose.^16^

### Residual-based detrending

Here, we explore this technique using the temperature, humidity and precipitation data. This methodology has more commonly been used for temperature, but it can be used for other climate variables such as precipitation and humidity, which are often highly influential to health outcomes. Since precipitation and humidity are physically linked to temperature and the smoothed GMTA can be used as a proxy forcing variable for regression, assuming the main anthropogenic forcing signal is captured by temperature change. The method is particularly attractive in the absence of complex climate models or computational resources. Furthermore, this approach may be more appropriate for precipitation and humidity data, as these variables tend to have more complex and spatially heterogeneous patterns and are less likely to possess a long-term trend, thus making data-driven detrending more challenging.^20^

Using the synthetic monthly long-term climate variables (temperature, humidity and precipitation) and the time-series of global GMTA values, we harmonise these into one data frame to create a counterfactual climate for each variable.^17,18^ Raw GMTAs include substantial interannual variability which is not directly attributable to human-forcing. Therefore, the first step is to smooth GMTAs with a smoothing method (here we used SSA) to capture the low-frequency forced climate signal, reduces internal variability, stabilises attribution coefficients, avoids arbitrary smoothing choices, and yield more robust counterfactual reconstructions that are only the long-term changes which are most likely attributable to human-influence on the climate system.^3,14^

Once the GMTA are smoothed using SSA,^20^ the linear relationship can be modelled and extracted to create the counterfactual climate variable. The fitted values from the regression (i.e. the part of the local climate variable that is linearly explained by GMTA) is subtracted from the observed data. That subtraction leaves the residuals, which are interpreted as the counterfactual climate variable — i.e. what the climate might have looked like without the influence of global warming. When using monthly data, the regression needs to be performed in separate months (run 12 independent regressions). If the regression is carried out over the entire dataset without accounting for month, the model could misinterpret seasonal variation (e.g., winter vs summer temperatures) as part of the trend. Regressing month-by-month allows the removal of the trend while preserving seasonal structure.^14,17,18^ Generating a climate counterfactual in this way is expressed mathematically below,

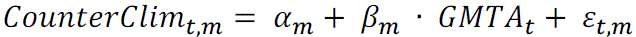

where, *t* is the indexed time (year), *m* corresponds to the month in each independent regression and therefore, α*_m_* is the month specific intercepts, β*_m_* · *GMTA_t_* is the fitted trend / slopes (the climate response to global warming) and ε*_t,m_* are the residuals, which define the counterfactual climate variable (*CounterClim_t,m_*). Figure 2 shows the observed and counterfactual climate variables using the residual-detrending method outlined here. The same methodology could also be applied to non-linear detrending, by swapping out the linear term with a non-linear smoother, such as a loess or spline.^17^

**Figure 2.**
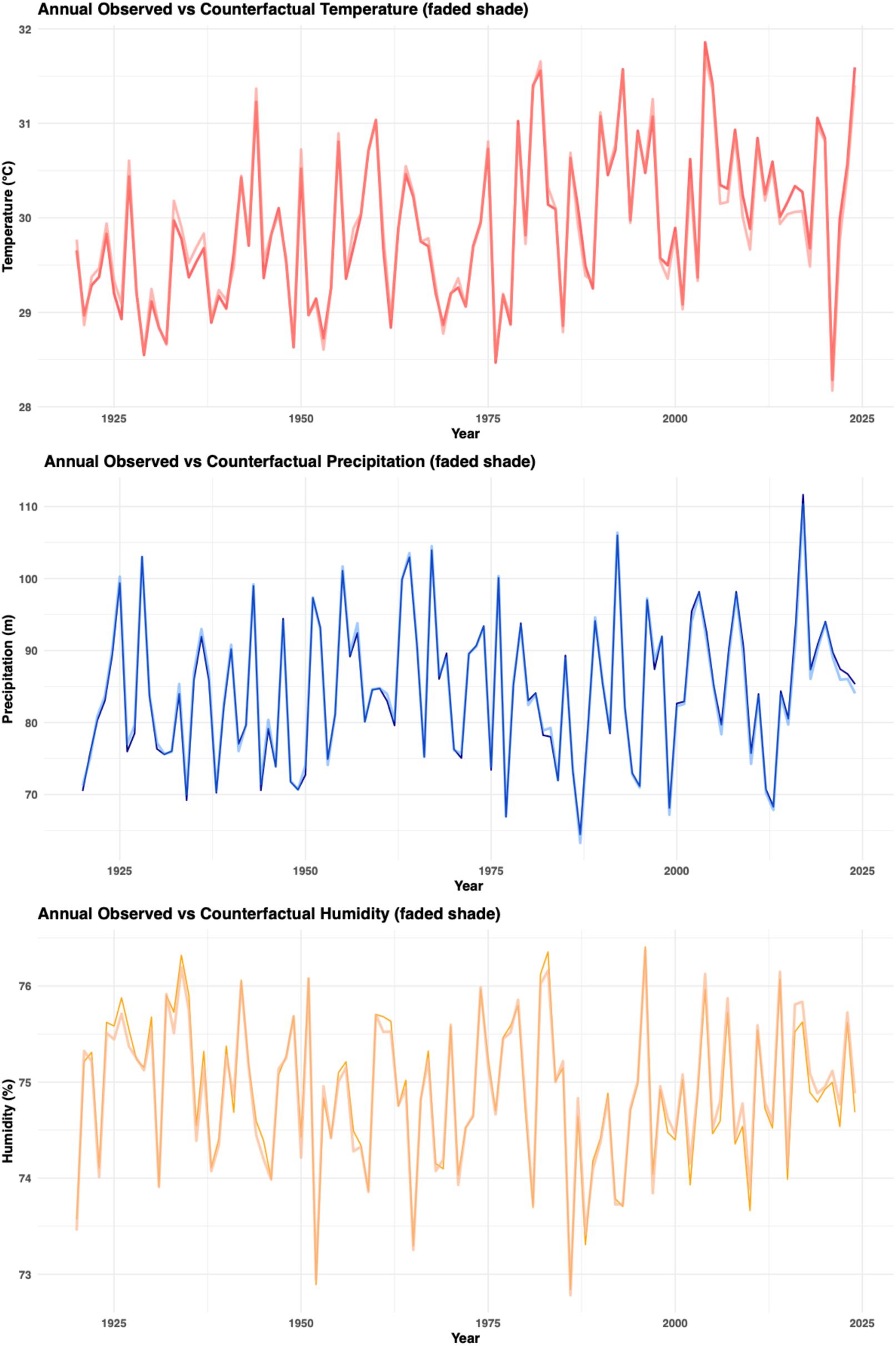
Annual averages of the monthly observed temperature, precipitation and humidity data and their corresponding counterfactual according to the residual-based detrending method. More pronounced differences between the counterfactual and observed would be expected over a longer observation period and at higher latitudes.

Using a predictive health model and/or dose-response function, attribution can be expressed in two ways: as absolute attribution — the difference in predicted health outcomes between the observed and counterfactual climates in the unit of the health outcome — or as fractional attribution — this difference expressed as a proportion of the total burden. Potential ways to communicate these results would include: “X% of the observed health impact over the study period is attributable to the observed trend, which is largely driven by anthropogenic forcing.” or “Our results suggest that climate change altered the health outcome by X over the study period.”.

It is important to remember that this is conditional attribution — that is, given that the health model and all other variables are held constant, the estimated difference in health outcomes is attributable to the observed climate trend, which is largely driven by anthropogenic forcing, rather than to anthropogenic climate change in its entirety. This conditionality also means that attribution here reflects only the temperature-mediated pathway of climate change, rather than the full suite of climate variables that may influence the health outcome. Furthermore, these methods cannot be used to assign a difference in a health outcome to a specific climate emitter or to a given level of greenhouse gas emissions, as emissions or concentration pathways were not used as the forcing term.

### Data-driven decomposition detrending

Using a data-driven decomposition method may be particularly desirable if you believe your climate variable in your location of interest may have a more complex climate that is not well represented by the global GMTA. There are multiple decomposition methods available, and in the tutorial materials there is a full explanation of both SSA and EMD, including how to set their parameters appropriately for your data. Despite methodological nuances, both methods follow the same principle, which is that they aim to decompose complex, nonlinear, or nonstationary time series into simpler, interpretable components. The methods break down a time-series into a set of components to reveal the trends (slow varying parts), oscillatory modes (cyclical or periodic patterns) and internal variability of the climate system (high-frequency residuals).^16^

SSA is better for long time-series and wide geographic areas, as these tend to have a clearer trend that can be removed,^20^ whereas, EMD is efficient when using fewer data points (e.g., smaller spatial or temporal resolution or scale) with more complex and non-linear relationships.^21^ The example provided in the tutorial materials uses the temperature data and decomposes it using SSA (Figure 3) to ease computational time (EMD is more computationally intensive). First the SSA object is created, being mindful of the window length (L), which is set depending on the temporal granularity of the data and to capture oscillations in the system at different time scales. Next, the eigenvalues are plotted in a scree plot, where usually the first two components should have high values (the trend), followed by a sharp decrease for the seasonality and noise. The time-series is then reconstructed, with only the first 2 components (the trend) and this is used in the same way as the linear model residuals above, subtracting these from the observational weather data to create the counterfactual.^20^

**Figure 3.**
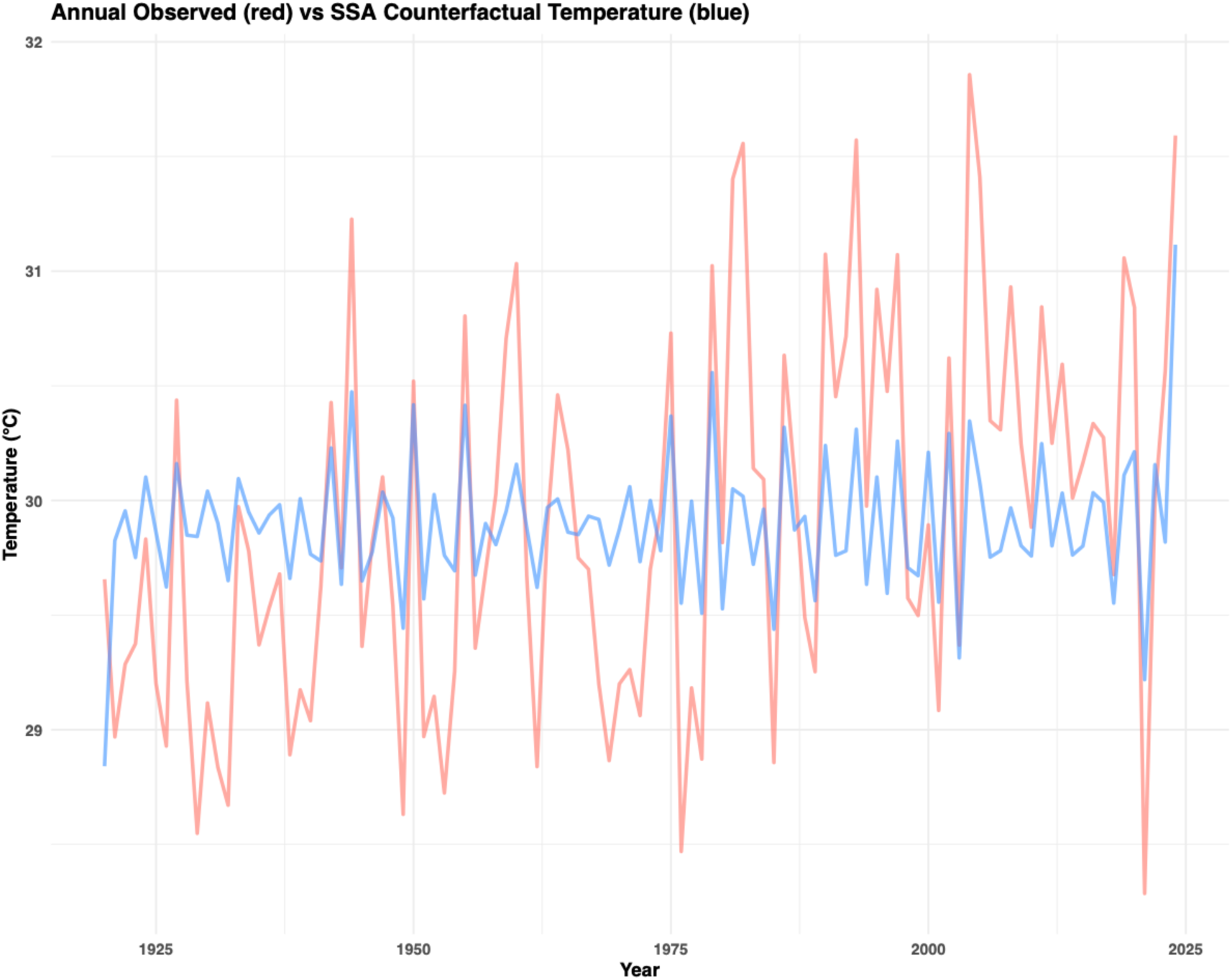
Annual averages of the monthly observed temperature data and its corresponding counterfactual according to the data-driven decomposition method, Singular Spectrum Analysis (SSA).

### Incorporation of GHG concentrations

In the methods that have been explored so far attribution can only be made between the health outcome and global warming, not specific concentrations of greenhouse gases e.g., CO₂, CH₄, N₂O, O₃, because the forced component was temperature variability (GMTA). However, if attributing to greenhouse gases is the aim of the study, then GMTA can be swapped out of the linear residual-based approach and replaced with concentrations data. Only one emitter should be explored at once (or GMTA) as these are highly correlated and would make the β values hard to interpret.^23^ Concentrations define the levels of greenhouse gases in the atmosphere (given in parts per million) and therefore provide information related to climate forcing. Modern greenhouse gas concentration datasets such as the NOAA Mauna Loa data are not available for pre-industrial times, however, these can be combined with paleoclimate records, such as the Law Dome ice cores.^24,25^ The example in the tutorial includes the long-term monthly temperature data, with the exploration of concentrations of atmospheric CO₂ as the forcing components in the models. Using these counterfactuals in HIA analyses, the following statements could be made: “A X% increase in [health outcome] over the study period is attributable to the observed increase in atmospheric GHG concentrations” or “A X% change in [health outcome] over the study period is attributable to rising anthropogenic CO₂ emissions.”.

## Counterfactual climate events

So far the methods explored here have only covered trend-based climate counterfactuals, but event-based attribution for HIA is also a growing field, particularly attributing excess mortality to heatwaves.^6,8^ Exploring new methods to create event counterfactuals are particularly important, as specific lived events may not be available in free running GCM datasets such as ISIMIP or DAMIP repositories. One or multiple extreme events can be related to several communicable and non-communicable health outcomes and how the counterfactual is generated depends on the aim of the HIA study and the trade-off between simplicity and how much physical realism is imposed. For example, if the aim is to explore the impact on the health outcome of a given event in a 2°C warmer world, greater physical realisms would be needed, however, if the aim is to explore the relationships between the health outcome and different components of the extreme event (e.g., temperature, precipitation, humidity), then a simple sensitivity analysis approach (manually altering the components of the event) could be used. In event counterfactuals there are two pathways: (1), a world “without the event”, which can include a trend-style approach e.g., removing the trend outlier that relates to the event and (2), a world with a “slightly different event” e.g., a cyclone with 10% faster wind speed, a more slow moving event, an event which takes a different track etc and compare these to the health outcome of interest.

The counterfactual approaches described here are not designed to formally attribute changes in health outcomes to anthropogenic climate change via an extreme weather event — that is, to claim that climate change made event X this much worse, which in turn caused Y change in the health outcome. Rather, these methods explore how variations in the characteristics of extreme events — as constructed in counterfactual worlds — propagate through to health outcomes. Attribution of the health impact can therefore be made to the extreme event itself but connecting that to anthropogenic climate change would require an additional, separate event attribution step. While such end-to-end attribution may become feasible as methods mature, we do not consider it methodologically robust to embed that link within a health impact assessment at this stage. Discussion of how the extreme events used here relate to anthropogenic climate change is therefore reserved for more descriptive analysis.

### “World without the event” counterfactuals

The simplest method to assess the impacts of an extreme event on a health outcome is to simply remove the outlier which corresponds to the extreme event e.g., the heatwave, storm etc. Fitting the climate-health model with and without the inclusion of the outlier and comparing the results to attribute its impacts.^40^ Two methods are explored in the tutorial to remove the heatwave events from the temperature data, the first being Fourier decomposition, which fits sine and cosine waves to describe climate data by its dominant periodic signal (great for periodic signals, but not for abrupt seasonal transitions), essentially the method captures trends and seasonal patterns which then allows the removal of anomalies like extreme events.^29^ The second method uses a simple time interpolation (linear interpolation which is better for short-term events), and essentially removes the event by drawing a straight line between the observation before and after the event. Figure 4 shows a comparison of the results for these two methods. In these types of “no-event” analyses, communication of the results could be as follows: “In the absence of the [event], we estimate X fewer hospital admissions (95% CI: Y–Z) would have occurred in [location] over [time-period]. This represents a Q% reduction compared to the observed burden.” or “Our results suggest that the [event] contributed approximately R% of the total observed health outcome during the study period, when compared to a physically plausible counterfactual scenario without the event.”

**Figure 4.**
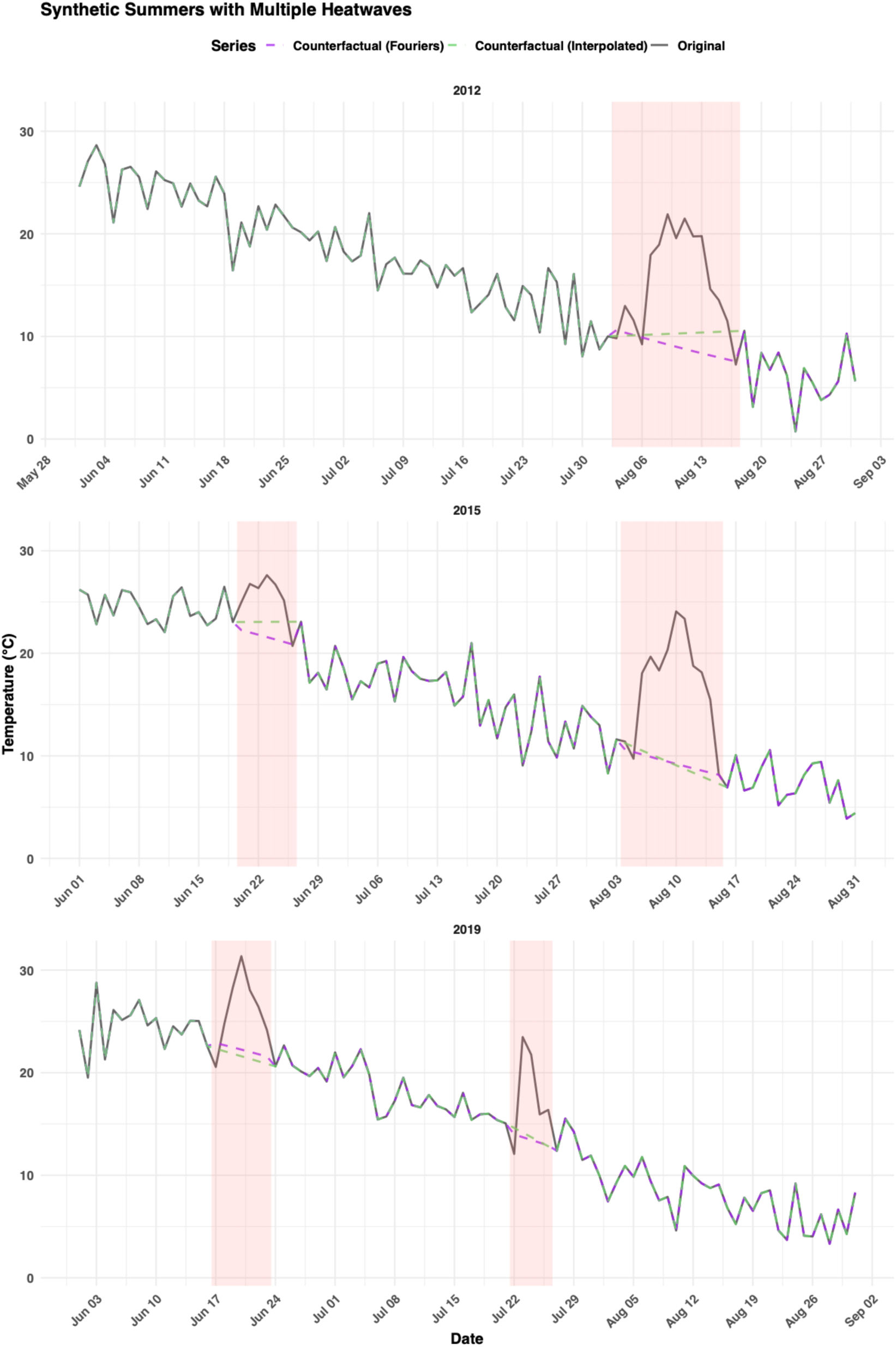
Observed temperatures (black line) and heatwaves (red shading) over multiple summers during the simulation period (Jun-August), 2012, 2015 and 2019, with the linear interpolation and Fouriers temperature counterfactuals during the heatwave periods.

### “World with different event” counterfactuals

There are several methods that could be used to simulate a slightly different event, and this will depend on the aims of the HIA analysis and a trade-off between interpretability/simplicity and physical realism. The first and perhaps most simple approach would be to adopt a qualitative storyline, which involves manually changing the parameters of the event to better understand how different elements of the event affected the health outcome e.g., 10% windier, 20% hotter etc.^41^ Here, the interest is less about the effect on the health outcome in real world events, but rather an exploratory exercise of the dose-response relationship. The approach would essentially be a sensitivity analysis of the event/events of interest and may be useful if only one past extreme event is available/of interest and therefore less data to build a more quantitative model. In the tutorial example, we explore creating counterfactual temperatures for the synthetic heatwave data that becomes 2% hotter, first converting to an absolute scale (Kelvin) and then increasing temperatures by 2%.

The second method of developing the counterfactual climate variable is based on known empirical relationships in atmospheric thermodynamics. These are global estimates and how weather translates regionally can be very different, therefore this is perhaps a better approach for larger spatial scales. Some important and useful formulas for generating a counterfactual include:

● Clausius-Clapeyron Relation = 1°C warming increases atmospheric moisture by 7%.^30^
● Precipitation Scaling with Temperature = 1°C warming increases precipitation by ∼2-3% (slower than water vapor because it is limited by energy balance. However, extreme precipitation scales closer to ∼6-10% per °C, particularly for convective storms.^31,32^
● Convective Available Potential Energy (CAPE) = 1°C warming increases convective energy by ∼10%, amplifying convective extremes.^33,34^

The example in the tutorial creates +2°C counterfactual temperature data for the same heatwave data (temperature), and using the outlined relationships above: water vapor, precipitation and CAPE were synthesised from the increase in temperature using the above scaling.^30,31,33^ Humidity values were then generated from temperature, water vapour and pressure, assuming surface pressure (1013.25 hPa). First saturation vapour pressure was calculated from temperature using the Tetens formula,^35^ followed by saturation specific humidity, actual specific humidity, actual vapour pressure and finally relative humidity. Combined these equations appear as below,

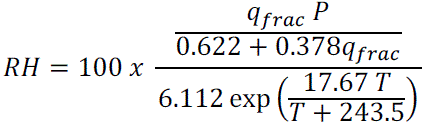

where

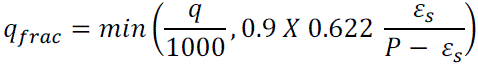

and

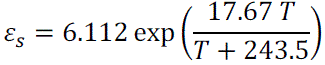

and *T* is air temperature (K), *P* is pressure (hPa), ε*_s_* is saturation vapour pressure (in hPa) and *q_frac_* is specific humidity (g/kg). All these relationships and the examples given would create counterfactuals in a warmer or cooler world for a variety of variables which are important for health.

Finally, we explore the use of stochastic weather generators (SWG) to generate multiple years of counterfactuals temperature data with heatwaves. A SWG is a statistical tool used to simulate long sequences of synthetic weather data (like rainfall, temperature, solar radiation, wind speed, etc.) that preserve the statistical properties of observed historical weather.^36^ Therefore, it can be used to simulate potential future events within the realms of possibility based on variability in historical events. In simple terms, it takes historical weather records (from a station or re-analysis) and learns the statistical patterns — such as how often it rains, how long wet/dry spells last, how daily temperatures vary, and correlations between variables. Then, it uses random sampling (stochastic methods) to generate new but realistic-looking weather time-series that can extend beyond the observed record.^37,38^ The models are frequently used in climate impact studies where long weather series are needed to test how a system responds to different climate conditions. It therefore is a good way of simulating future events but still maintaining physical realism. SWGs are useful because they can preserve long-term patterns in the weather but add the much-needed stochastic variation that is often lost in counterfactuals, particularly detrending exercises. They are also highly flexible and can be tweaked to whatever is needed, creating realistic alternative versions of an event to explore the health outcome.^39^

In the tutorial we use the synthetic heatwave temperature data and a generic ensemble is generated of plausible future daily temperature series with heatwaves (2020 to 2029), using Fourier decomposition (used above), single AR(1) and Gaussian noise to incorporate the stochasticity. Once the temperature data were loaded and the simulation parameters specified (warming trend, simulation window, heatwave threshold and number of simulations), the workflow proceeded in two modelling steps. First, a smooth seasonal baseline was estimated from the non-heatwave days using Fourier decomposition. Second, an AR(1) model was fitted to the residuals from this seasonal fit to represent short-term temporal dependence. Finally, 200 simulations were run incorporating stochastic variability, whereby each simulated value includes a random draw from a normal distribution with the estimated residual standard deviation, adding realistic day-to-day variability. These stochastic residuals were then added to the deterministic seasonal model to produce the full synthetic temperature series. When using multiple observational or simulation datasets, an ensemble with a confidence interval is recommended, and can be achieved via bootstrapping. The final stage of the tutorial takes the simulated heatwave data from the 200 simulations in the SWG and creates an ensemble value with confidence intervals for each data point in the simulation period.

It should be noted here that we are not attributing the extreme event to climate change, but rather the extreme event (or components of the extreme event, which may have been altered by anthropogenic climate change) to the health outcome. In theory, you could use the empirical relationships and work the other way to generate a pre-industrial counterfactual (e.g., 1.5°C cooler) for the event. Additionally, you could apply any of the data-driven decomposition methods to some extreme event data (e.g., heatwaves), incorporating them into long-term climate data and then removing the trend to generate a pre-industrial event counterfactual.

## Conclusion

This tutorial provides a robust, flexible and accessible framework for developing counterfactual climate variables, particularly for impact-based analyses where climate models or High-Performance Computing may not be available. We have explored several methodologies and while we hope some users may use our tutorial to deepen their understanding, we anticipate others may have specific climate data they want to explore counterfactuals for and Figure 5 provides a flow diagram to guide readers to the correct method for them.

**Figure 5.**
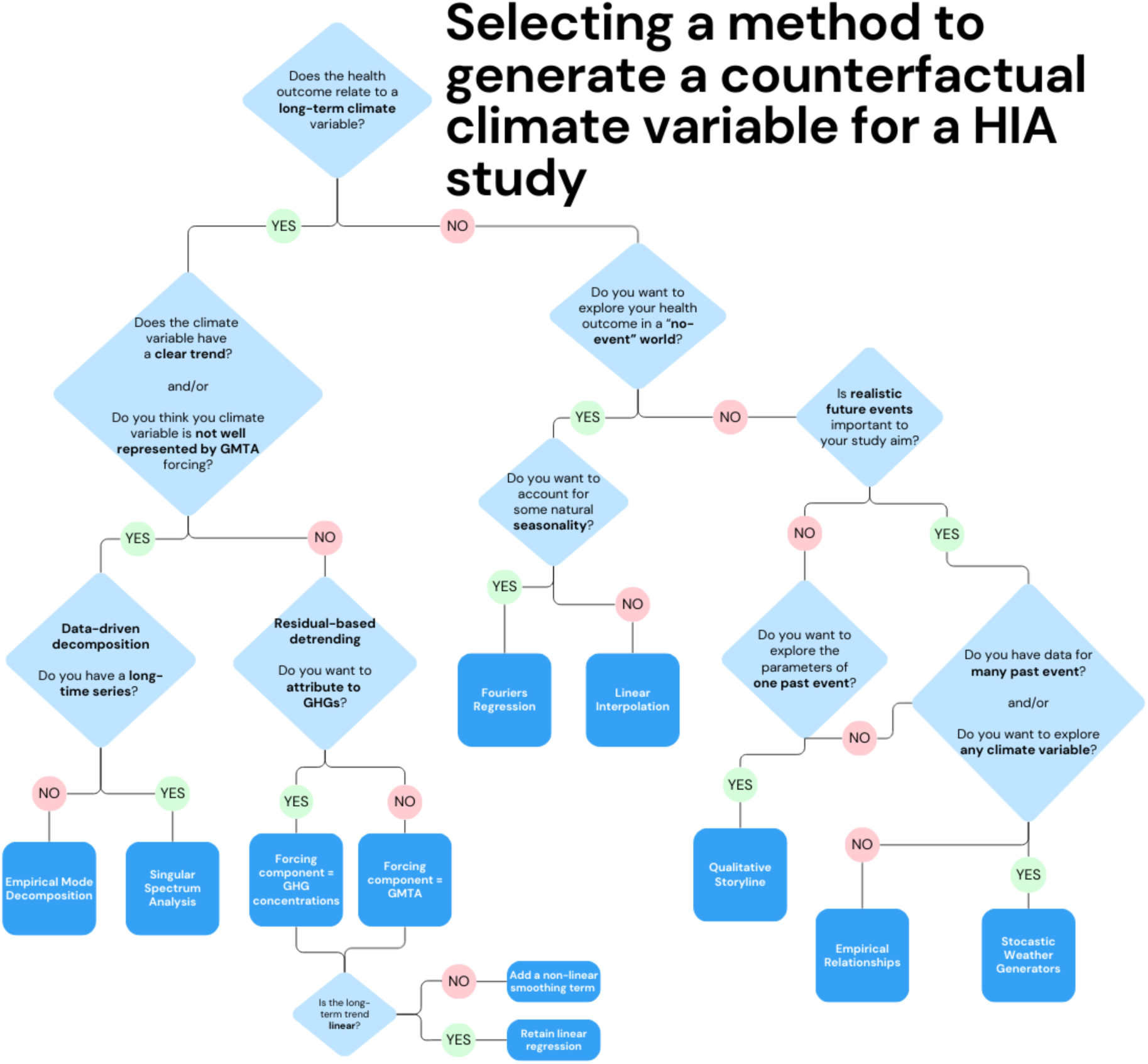
Once a robust health-exposure model has been developed, the flow diagram suggests a range of decisions to select the correct method to generate the counterfactual climate variables needed. *Source*: Author’s own

While global circulation models remain the gold standard for generating counterfactual climates, this tutorial offers a practical alternative for researchers in health impact assessments and related attribution studies with limited access to running bespoke counterfactual climates via GCMs. It highlights ATTRICI-style methods and demonstrates how it can be applied across diverse contexts. We hope the manuscript, along with the accompanying R tutorial and repository, helps build capacity in the HIA community, improving accessibility, interpretation, and collaboration between the climate and health sector.

## Code & Data Availability

All code is available in the GitHub repository link below; this includes the tutorial and all data generation and processing scripts: https://github.com/GinaCharnley/Developing-Counterfactuals-for-HIA

## Data Availability

All code is available in the GitHub repository link below, this includes the tutorial and all data generation and processing scripts

https://github.com/GinaCharnley/Developing-Counterfactuals-for-HIA

